# Berrylyzer-an Efficient, Traceable, and Lightweight Intelligent Agentic System for Prenatal Genetic Diagnosis

**DOI:** 10.64898/2026.04.02.26349929

**Authors:** Meng Meng, Ling Liu, Qianqian Du, Xinyao Zhou, Yuan Tian, Kuan Sun, Naiqi Li, Pinggui Zhang, Xianyi Lian, Ning Fan, Na Zhu, Shujing Li, Aiping Mao, Yuezhen Li, Gang Zou

## Abstract

**Background:** Artificial intelligence (AI)-driven variant prioritization has demonstrated substantial utility in expediting genetic diagnosis by ranking the most likely causative variants. While a variety of tools have been developed, few address the unique clinical and technical constraints in prenatal genetic diagnosis.

**Methods:** We introduce Berrylyzer, a novel, end-to-end variant prioritization system applied to prenatal diagnosis. Inspired by clinician’s reasoning process during variant interpretation, Berrylyzer applies a modular, stepwise scoring architecture that jointly integrates phenotypic and genomic evidence and delivers a ranked list of candidate variants, achieving high computational efficiency without compromising analytical rigor. Moreover, Berrylyzer natively supports both structured ontologies and free-text clinical narratives, enabling flexible integration into diverse clinical environments. Its performance was rigorously evaluated across two independent, real-world prenatal cohorts and benchmarked against three state-of-the-art methods: Xrare, Exomiser, and PhenIX.

**Results:** Across the two datasets, Berrylyzer ranked 56.41% and 58.12% of diagnostic variants first, and achieved recall rates of 94.02% and 97.42% within top 20, respectively. Berrylyzer outperformed Xrare (85.19% and 87.08%), Exomiser (84.90% and 85.98%), and PhenIX (82.05% and 88.93%). Stratified analysis consistently demonstrated superior performance across diverse disease categories, inheritance patterns, and analytical strategies. Notably, Berrylyzer exhibited robustness regardless of phenotype forms, yielding comparable top 20 recall rates for free-text descriptions and standardized terminologies.

**Conclusion:** Berrylyzer represents an accurate, interpretable, and computationally lightweight variant prioritization system for prenatal genetic diagnosis. The superior performance across heterogeneous diagnostic contexts enables it as a practical solution for seamless integration into clinical pipelines, thereby advancing precision medicine in prenatal settings.

## Introduction

Congenital disorders, defined as structural or functional anomalies occurring during intrauterine development, represent a significant global health burden, affecting approximately 6% of newborns worldwide and contributing to over 200,000 neonatal deaths annually according to the World Health Organization (1). Accurate and timely prenatal genetic diagnosis serves as the cornerstone for evidence-based parental counseling, reproductive decision-making, and obstetric and perinatal management. The advent of sequencing technologies has revolutionized the diagnostic landscape of prenatal medicine, substantially enhancing diagnostic yield and expanding the spectrum of detectable genetic conditions. However, identifying the causative variants associated with fetal phenotypes remains extraordinarily challenging. Whole-exome sequencing (WES), now widely adopted in clinical practice, typically generates tens of thousands of variants per individual case (2). While conventional bioinformatics filtering pipelines can reduce this overwhelming volume to several hundreds of candidate variants, subsequent manual curation and interpretation of these candidates to find 1∼2 causative variants remain time-consuming and labor-intensive, and highly rely on specialized expertise in clinical genetics. This bottleneck for clinical interpretation is particularly acute in prenatal settings, where decision-making windows are compressed. Thus, there exists an urgent need for robust and clinically deployable tools capable of prioritization of candidate variants, thereby reducing interpretation burden, compressing diagnostic turnaround time, and supporting accurate prenatal genetic diagnosis.

Artificial intelligence (AI) is rapidly revolutionizing the world of clinical genetics and molecular medicine, as in many other fields (3). AI-driven variant prioritization methods have been developed to prioritize the most likely causative variants associated with patient’s phenotypes. Typically, these variant prioritization tools can be classified into “genotype-driven” approaches which exclusively leverage genotype information (such as MutationTaster (4) and CADD (5)) and “phenotype-driven” approaches which integrate both phenotype and genotype information. Empirical evidence consistently demonstrates that “phenotype-driven” approaches achieve superior accuracy and have become the prevailing paradigm (6). Over the past two decades, numerous new variant prioritization tools are blooming (7–10). Exomiser, the most widely adopted phenotype-driven variant prioritization tool, has been applied in the primary diagnostic pipelines in some large-scale genome projects, such as the 100,000 Genomes Project and the Undiagnosed Disease Programme (11–13). Despite these advances, existing tools present several critical limitations. First, although widely employed in pediatric and adult genetic diagnosis (14, 15), there exists no prenatally oriented variant prioritization tool to date. This gap is largely due to the limited knowledge of prenatal manifestations of Mendelian disorders in major public disease databases, such as the Online Mendelian Inheritance in Man (OMIM) which only includes explicit description of prenatal manifestations in 460 disease entries now. Moreover, the dynamic, evolutional nature of fetal phenotypes across gestation complicates the situation (16). Second, most variant prioritization tools are designed with Human Phenotype Ontology (HPO) as a consensus input vocabulary; consequently, clinicians are required to engage in labor-intensive manual extraction or deploy auxiliary computational tools, such as ClinPhen (17), which introduce delays and dependency on additional tools. To scale with the booming demand for clinical genetic diagnosis, variant prioritization tools need to be improved to handle free-text phenotypic terms in medical records. In recent years, large language models (LLMs) have shown impressive capabilities in parsing structured and unstructured data, motivating the development of end-to-end prioritization frameworks, such as Genetic Transformer (18) and DeepRare (19). However, LLMs remain hindered by two persistent challenges: hallucination which refers to factually incorrect, irrelevant, or inconsistent results (20) and architectural opacity (“black-box” behavior). These issues undermine the prediction reliability and limit practical application. Meanwhile, the trillion-parameter scale of state-of-the-art LLMs generates significant practical barriers to integration and computational efficiency in real-world clinical environments.

To address these unmet clinical and technical challenges in prenatal diagnosis, we introduced Berrylyzer, a novel, clinically validated AI framework for prioritization of candidate variants of Mendelian disorders in prenatal settings. Berrylyzer presents four key characteristics that collectively advance accuracy, interpretability, and deployment feasibility:

(1) an automated end-to-end framework: Berrylyzer supports heterogeneous phenotypic inputs, presented as either free-text descriptions or standardized HPO terms, thereby eliminating the HPO curation bottleneck while maintaining compatibility with established clinical workflows;
(2) vocabulary of prenatal manifestations: the framework incorporates a comprehensive, rigorously curated dictionary of fetal phenotypes, enabling precise mapping between observed prenatal features and established diseases and genes;
(3) stepwise scoring architecture: in addition to final outputs, Berrylyzer also offers stepwise, intermediate results, facilitating analytical traceability and clinical validation;
(4) computational efficiency and lightweight architecture: Berrylyzer is engineered upon efficient algorithms, allowing for rapid and accurate variant prioritization with low computational resource consumption, thereby enabling feasibility of deployment in clinical practice.

Berrylyzer was rigorously and comprehensively evaluated based on two independent, real-world prenatal cohorts. Through benchmarking against three state-of-the-art tools (Exomiser, Xrare, and PhenIX), we demonstrated the robustness of Berrylyzer in prioritization accuracy and yield across diverse diagnostic contexts. Collectively, these findings establish Berrylyzer as an accurate, traceable, and lightweight strategy capable of accelerating variant interpretation of prenatal genetic diagnosis, thereby advancing prenatal precision medicine.

## Materials and methods

### Pipeline overview

Berrylyzer is an AI-driven framework designed to emulate the expert reasoning process during variant interpretation and prioritize candidate variants that are most likely responsible for observed phenotypes. It receives multi-model input data, including genome sequencing data stored in variant call format (VCF) files and clinical phenotypes presented as either free-text clinical narratives or standardized HPO terms. When multi-sample VCF files from family members are included for the analysis, a corresponding pedigree file must also be provided. Subsequently, by integrating two parallel, domain-specific modules: one dedicated to phenotype matching and the other to variant featuring, it outputs a ranked list of candidate variants (Figure 1A). Expert clinicians can review the top-ranked variants for diagnostic confirmation and final reporting.

**Figure 1.**
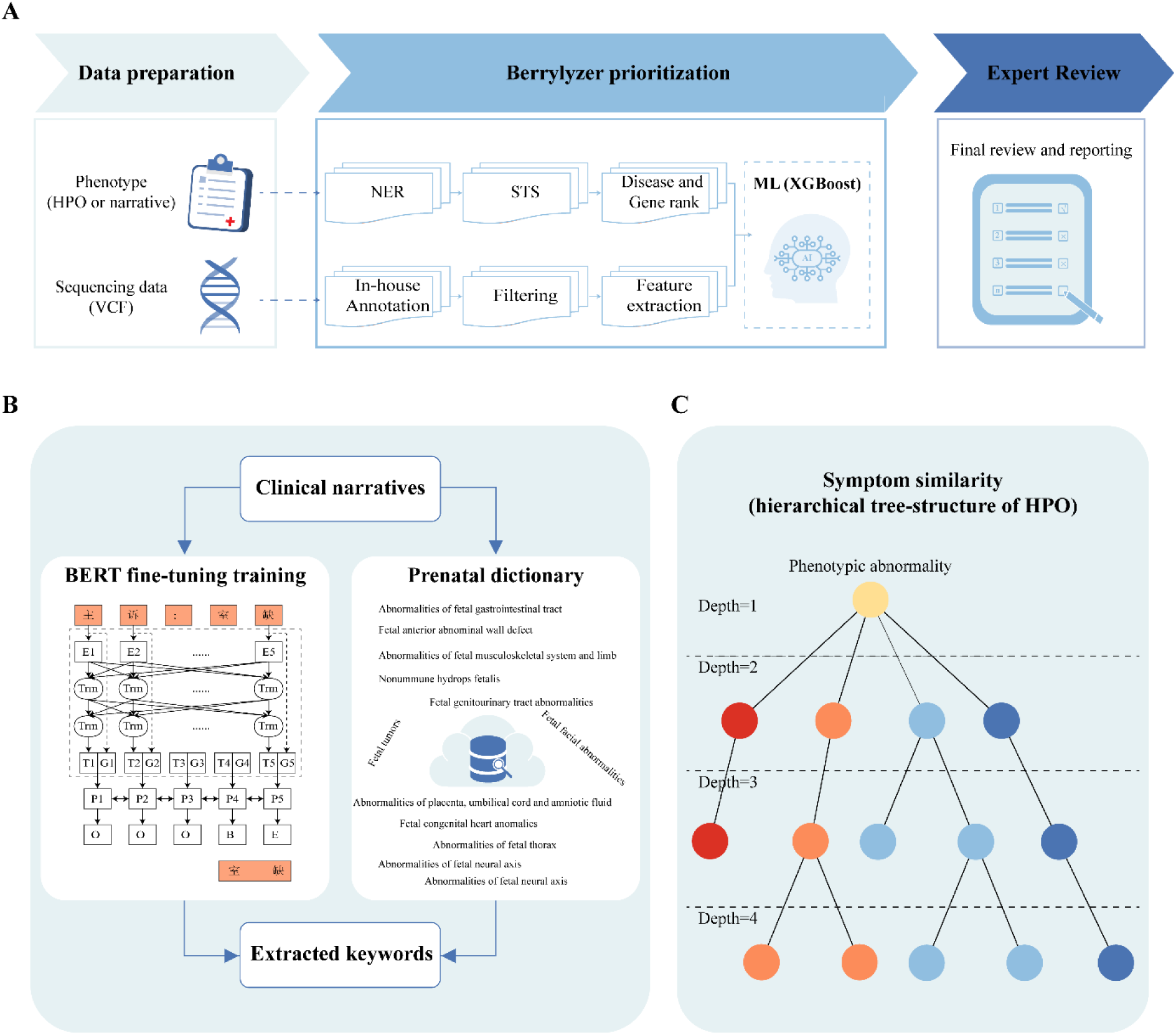
Illustration of the pipeline of Berrylyzer. (A) Overview of Berrylyzer workflow. Berrylyzer accepts multi-modal input data, including sequencing dada provided in the form of VCF file and clinical phenotypes in various forms, such as free-text descriptions, HPO terms, and disease names. The phenotypic information is extracted for keywords, standardized based on the semantic similarity between observed manifestations and phenotypic profiles of known human diseases, and accordingly scored at disease-as well as gene-levels. Parallelly, an annotation and filtering process are assigned to each variant to filter high-confidence candidates and provide evidence for subsequent feature engineering. By integrating features from phenotypic and genomic evidence, Berrylyzer outputs a ranked list of candidate variants for curation by expert clinicians, thereby confirming the diagnosis. (B) The schematic diagram of dual-matching protocol for keywords extraction, comprising named entity recognition base on NLP technology and identification through construction of prenatal dictionary. (C) Illustration of the hierarchical tree-structure of HPO terms.

### Phenotypic scoring module

The phenotypic scoring module computes the semantic similarity between user-provided clinical phenotypes and phenotypic profiles of certain diseases.

#### (1) Keywords extraction

To overcome the scarcity of well-annotated fetal phenotypes in current disease databases, an innovative dual-matching protocol was implemented (Figure 1B). First, a pre-trained Chinese Bidirectional Encoder Representation from Transformers (BERT) language model (21) was employed for named entity recognition (NER) based on natural language processing technology (NLP), which extracted phenotype-relevant keywords from unstructured clinical narratives. To ensure robustness and generalizability, a large-scale, de-identified in-house corpus of prenatal clinical notes was collected and rigorously annotated by clinical geneticists; these high-quality annotations served as the gold-standard training set for supervised fine-tuning of the BERT model. Second, recognizing the limited coverage of prenatal manifestations in current disease databases, we constructed a comprehensive, manually curated dictionary of abnormal prenatal manifestations. This dictionary synthesized evidence from two authoritative clinical references, including “Prenatal Ultrasonographic Diagnosis of Fetal Abnormalities” and “Callen’s Ultrasonography in Obstetrics and Gynecology”, and aligned entries with HPO framework. Notably, the HPO community is now giving effort for the expansion of prenatal phenotypes to support precision prenatal genetic diagnosis (16). Thus, our dictionary enables systematic mapping of prenatal findings to HPO terms, and by extension, to associated diseases in OMIM database through disease annotation in HPO database. In addition, it also incorporated commonly used clinical abbreviation, Chinese translations of disease names in OMIM database, English acronyms, and gene symbols. Outputs from both NER-driven extraction and dictionary-based matching were jointly integrated to produce the final keywords. Collectively, this dual-matching strategy enhances both accuracy and prenatal adaptability in phenotype keywords identification.

#### (2) Keywords standardization

A fine-tuning strategy was adopted for keywords standardization through vector representation and cosine similarity calculation. Specifically, supervised training was performed to learn embedding that captured the hierarchical relationship within the Chinese HPO (CHPO) tree structure, based on the theory that CHPO terms sharing proximal ancestor nodes exhibited higher similarity (Figure 1C). Formally, if node x represents the deepest common ancestor of node i and j in the CHPO hierarchy, their theoretical semantic similarity is calculated as follows:

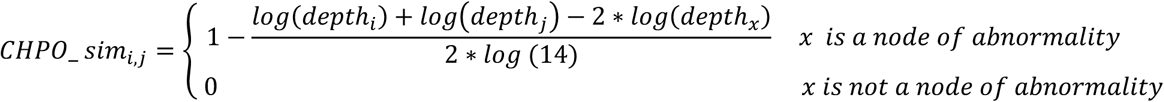

Subsequently, expert clinicians manually mapped the extracted keywords to standardized CHPO terms, forming a high-fidelity supervised dataset for learning similarity metrics between free-text descriptions and ontology terms. In addition, disease names and clinical synopsis of diseases in OMIM database also were adopted as standardized ontologies, further improving the generalizability of standardization.

#### (3) Disease and gene scoring

Berrylyzer leveraged a pre-compiled, bi-directionally curated candidate gene and disease knowledge base derived from the OMIM database, wherein only genes with definitive disease associations and diseases with definitive causative genes were retained. For a given disease *i* associated with *m* CHPO phenotypes, and *n* extracted phenotype keywords, the similarity matrix for the disease was:

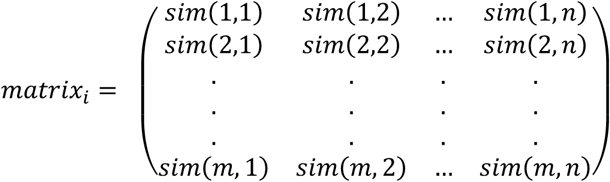

Then, the disease-level score was computed as:

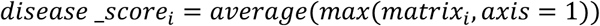

If a gene *x* was corresponded to n diseases, then the gene-level score was computed as:

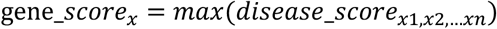

Gene-level scores were normalized to [0,1], and then used for ranking of candidate genes.

### Variant annotation and filtering module

The first step of this module was to comprehensively annotate each variant using an in-house, clinically validated bioinformatics pipeline. These annotations described the genomic location, transcript, variant type, information in variant databases (HGMD and ClinVar), *in silico* predictions, minor allele frequency (MAF), inheritance pattern, corresponding gene and disease, and notably a streamlined classification level and evidence based on guideline of the American College of Medical Genetics and Genomics and the Association for Molecular Pathology (ACMG/AMP) (22) through an in-house Bayesian classification framework. Subsequently, variants were filtered according to the following criteria: (1) MAF exceeding a predetermined threshold; (2) classification as benign and likely benign under the Bayesian classification framework; and (3) on genes absent from the precompiled gene-disease knowledgebase. The following variants were retained: (1) variants in the whitelist which was curated based on previously diagnosed cases; (2) variants which were annotated as pathogenicity/likely pathogenicity in ClinVar or HGMD.

### Variant prioritization

To embed domain-specific genetic and clinical expertise into Berrylyzer’s decision logic, seven groups of biologically and clinically interpretable features were selected for supervised learning: (1) features of associated gene; (2) evidence according to the ACMG/AMP guideline; (3) sequencing quality metrics; (4) inheritance pattern; (5) population frequency; (6) annotation in variant databases; *and* (7) *in silico* predictions. The XGBoost model (23) was trained to assign each variant a final score ranging from 0 and 1 for prioritization. Training was performed based on in-house, de-identified dataset including clinical phenotypes, VCF files, and variants in clinical reports.

Berrylyzer provided not only the ranked variant list but also transparent, stepwise intermediate outputs to support clinician review for final report. These include: (1) extracted keywords that can be manually reviewed and refined; (2) standardized CHPO and HPO terms with corresponding identifiers; (3) disease-level score and ranking; (4) gene-level score and ranking; (5) ACMG/AMP classification level and supporting evidence; and (6) functional annotations, such as variant type. This granular transparency enables clinicians to audit, refine, and validate analytical decisions in accordance with institutional protocols and professional judgment.

### Application

Berrylyzer is accessible as a secure, interactive web server (https://www.berrycloudservice.com). For institutions requiring on-premises deployment, Berrylyzer is also available as a Docker container. The computational demand of the workflow is predominantly determined by the variant annotation module, which introduces a trade-off between runtime and memory consumption. Under the default configuration, processing a WES sample typically requires approximately 20 minutes and peaks at ∼10 GB of RAM. Alternatively, the high-performance configuration reduces runtime to within 10 minutes, albeit at the cost of elevated memory consumption (∼40 GB). This flexibility enables users to select the most appropriate setting according to their available computational resources. All other pipeline components are computationally lightweight, collectively requiring less than one minute to complete.

### Determination of benchmarking tools

The determination of benchmarking tools was performed according to criteria adapted from prior methodological studies (24): (1) phenotype-driven; (2) with active maintenance; (3) access to free, local installation to ensure data safety. Finally, three widely adopted tools were selected, including Exomiser (11), PhenIX (25), and Xrare (8). Exomiser v14.0.0 (with variant and phenotype databases version 2406) was downloaded from its official GitHub repository (https://github.com/exomiser/Exomiser/releases) and configured using default settings. PhenIX was available with Exomiser, and can be run in the same way as Exomiser. Xrare was deployed via its official docker image (https://web.stanford.edu/group/wonglab/Xrare/xrare-pub.2021.html) and invoked using the xrare R package. All tools were supplied with HPO terms, either manually curated by genetics expert or extracted from Berrylyzer.

### Benchmarking data collection

Multi-center prenatal cohorts were recruited in this study, consisting participants from The Shanghai First Maternity and Infant Hospital (TSFMIH) and The Third Affiliated Hospital of Zhengzhou University (TTAHZU). For each case, the de-identified data were collected, including VCF files, clinical phenotypes, and variants in clinical reports adjudicated by certified clinical geneticists. The reported variants included as follows: (1) diagnostic variants: pathogenic/likely pathogenic variants identified in a disease gene that partially or fully explains the fetal phenotypes; (2) potential variants: variants of unknown significance (VUS) identified in a disease gene that partially or fully explains the fetal phenotypes; and (3) secondary findings (SFs): pathogenic/likely pathogenic variants identified in ACMG recommended SF 3.2 gene list (26), irrespective of phenotypic relevance. Only single nucleotide variants (SNVs) and Insertions and Deletions (InDels) were included in this study.

### Performance evaluation

Clinically reported variants served as the “gold standard” for benchmarking. The performance was assessed using the top-K recall rate, specifically, the proportions of reported variants within top 1, top 2, top 3, top 10, and top 20 outputs for each method. In cases with more than one variants, each variant was separately evaluated.

## Results

### Overview of Berrylyzer

Figure 1 illustrated the end-to-end analytical workflow of Berrylyzer. Specifically, the pipeline started with uploading multi-modal input data, comprising phenotypic information presented either as free-text clinical narratives or structured HPO terms, and genomic data from the proband with optionally from other family members. Upon receiving inputs, the phenotypic scoring module implemented phenotypic keywords extraction, standardization, and further mapping to candidate human disease and genes in a precompiled knowledgebase; in parallel, the variant annotation and filtering module annotated each candidate variant and subsequently filtered high-confidence variants. Prioritization was achieved through integrative feature engineering that fused phenotypic relevance scores and variant-level evidence, yielding a composite prioritization score for each candidate variant. Berrylyzer provided not only the final ranked list of candidate variants, but also stepwise intermediate results, including extracted keywords, standardized phenotype terms, as well as disease- and gene-level rankings, ensuring a transparent and auditable interpretation process.

### Demographic characteristics of benchmarking participants

Participants were recruited from two independent prenatal diagnosis centers, forming the TSFMIH cohort (n=535) and the TTAHZU cohort (n=826). Table 1 presented the demographic characteristics of the participants. Among them, 87.29% underwent prenatal diagnosis due to the detection of fetal anomalies, while the remaining were tested due to others reasons, such as family history or history of adverse pregnancy. Each case in both cohorts had at least one variant reported. Cases with multiple variants included: (1) multiple variants in one gene reported to be diagnostic, potential, or secondary findings; (2) variants located on different alleles related to recessive disorders; and (3) variants found in different genes. These datasets reflected the complexity and diversity of real prenatal scenarios, ensuring alignment with real clinical practice.

**Table 1.** Demographic and clinical characteristics of the participants.

| Variables | TSFMIH | TTAHZU |
| --- | --- | --- |
| *Case number* | 535 | 826 |
| *Reason for prenatal diagnosis* |  |  |
| Fetal anomalies | 491 | 721 |
| Multiple anomalies | 110 | 195 |
| Musculoskeletal abnormalities | 70 | 123 |
| Neurologic abnormalities | 53 | 126 |
| Cardiovascular abnormalities | 65 | 91 |
| Genitourinary abnormalities | 32 | 67 |
| other abnormalities | 161 | 119 |
| Other reasons | 44 | 105 |
| *Analytical strategy* |  |  |
| Proband only | 37 | 212 |
| Trio | 498 | 614 |
| *Variant number* |  |  |
| Diagnostic | 351 | 542 |
| Potential | 101 | 587 |
| Secondary finding | 182 | 120 |

### Berrylyzer outperformed existing variant prioritization tools

To rigorously evaluate the performance of Berrylyzer, we conducted a comparative benchmarking analysis (Figure 2A) against three widely adopted variant prioritization tools selected for their high efficacy and free, locally programmable access: Exomiser (11), PhenIX (25), and Xrare (8).

**Figure 2.**
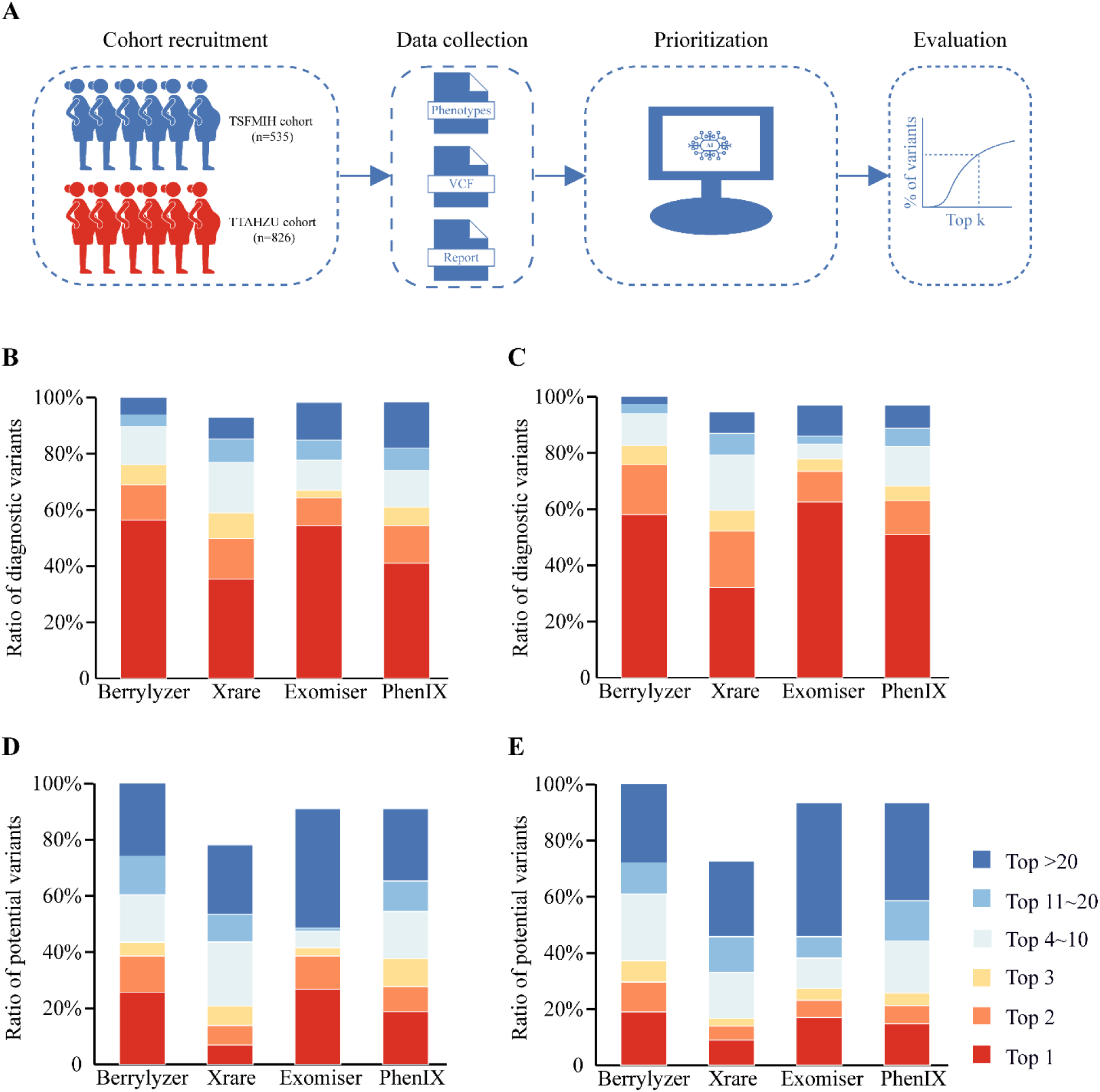
Overview of benchmarking workflows and performance evaluation results. (A) Diagram of benchmarking workflows in this study, from participant recruitment, data collection, method implementation, to benchmarking analysis. (B-C) Distribution charts of performance of Berrylyzer and other benchmarking methods for diagnostic variants on the TSFMIH (B) and TTAHZU (C) cohorts. (D-E) Distribution charts of performance of Berrylyzer and benchmarking methods for potential variants on the TSFMIH (D) and TTAHZU (E) cohorts.

Specifically, on the TSFMIH cohort, Berrylyzer accurately ranked 56.41%, 68.95%, 76.07%, 89.74%, and 94.02% of diagnostic variants within top 1, top 2, top 3, top 10, and top 20, respectively (Figure 2B). Notably, Berrylyzer assigned a rank to every variant although 1.14% of variants were ranked beyond top 100. In contrast, Xrare, Exomiser, and PhenIX only achieved recall rates within top 20 for 85.19%, 84.90%, and 82.05% of diagnostic variants, significantly lower than that of Berrylyzer. Furthermore, 7.12%, 1.71%, and 1.71% of variants remained unranked by these benchmarking tools, likely due to their restrictive filtration criteria. However, the majority of these variants were ranked within top 3 by Berrylyzer. Similarly, Berrylyzer also achieved an outstanding performance on the TTAHZU cohort, with a recall rate of 97.42% within top 20 (Figure 2C). When compared, the results showed that although Exomiser had slightly higher top 1 accuracy than Berrylyzer (62.55% *vs* 58.12%), it underperformed Berrylyzer at all subsequent ranks, and only achieved a recall rate of 85.98% within top 20. Xrare and PhenIX showed consistently lower performance across all ranks. Moreover, Xrare, Exomiser, and PhenIX failed to assign any rank for 5.35%, 2.95%, and 2.95% of variants, respectively, most of which were successfully ranked within top 3 by Berrylyzer. We further assessed the performance on potential variants (Figure 2D and E). As expected, Berrylyzer exhibited a reduced accuracy for these variants than diagnostic variants, which may due to the inherent biological and interpretive uncertainty associated with variants of uncertain significance. Nevertheless, Berrylyzer still significantly outperformed all other tools, underscoring its robustness across variant categories. Collectively, these results confirmed the robust performance of Berrylyzer across diverse datasets and reporting scenarios encountered in clinical practice.

### Ranking performance in different inheritance patterns

The prioritization task is more challenging for Mendelian recessive disorders than for dominant disorders, as pathogenic variants must be present in both copies of the gene investigated. Thus, we evaluated the performance of Berrylyzer across the autosomal dominant inheritance and autosomal recessive inheritance, respectively. For the evaluation, we combined diagnostic and potential variants together. In detail, Berrylyzer ranked 57.53% (Figure 3A) and 44.91% (Figure 3B) of autosomal dominant variants top 1 on the TSFMIH and TTAHZU cohorts, respectively, outperforming its performance for autosomal recessive variants (25.77% and 32.06%). Nevertheless, for both inheritance patterns, Berrylyzer achieved nearly identical accuracy when considering the rank within top 2, top 3, top 10, and top 20. In contrast, the three benchmarking methods consistently performed better on dominant cases than on recessive ones. When compared to benchmarking tools, Berrylyzer exhibited comparable accuracy on autosomal dominant variants to Exomiser across both cohorts and slightly higher than Xrare and PhenIX. For autosomal recessive variants, although Exomiser performed the best within top 1 (30.18% and 42.75%), Berrylyzer outperformed it within top 2 (49.56% and 58.02%), top 3 (58.59% and 64.89%), top 10 (79.07% and 83.21%), and top 20 (86.78% and 91.60%).

**Figure 3.**
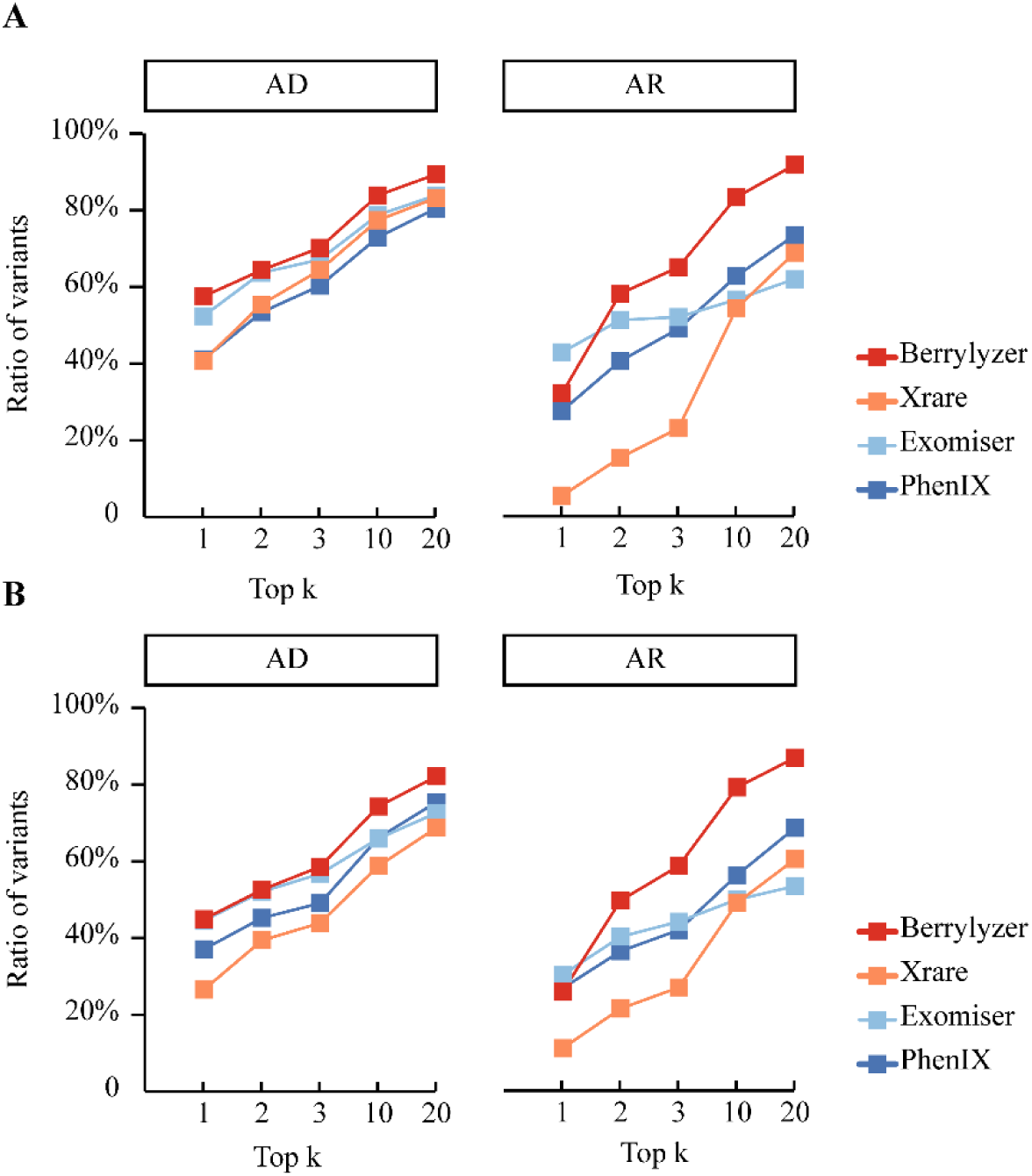
Distribution plots of performance for inheritance patterns. (A) Distribution plots of performance on the TSFMIH cohort; (B) Distribution plots of performance on the TTAHZU cohort. The distribution plots illustrate the ratio of variants ranked within top 1, top 2, top 3, top 10, and top 20 by each method. Each method is represented by a different color.

Additionally, we found that in some cases involving compound heterozygous variants of autosomal recessive diseases, Berrylyzer successfully ranked both variants, whereas three other benchmarking methods ranked only one. For instance, in one case, the fetus exhibited bilateral enlarged and echogenic kidneys along with oligohydramnios, leading to the clinical suspicion of infantile polycystic kidney disease. The diagnosis was confirmed as autosomal recessive polycystic kidney disease (ARPKD), caused by compound heterozygous variants in the *PKHD1* genes: c.9455del (pathogenic) and c.6332+40A>G (likely pathogenic). Berrylyzer ranked these two variants at top 1 and top 2, respectively. In contrast, Xrare, Exomiser, and PhenIX ranked only the former variant at top 1, top 60, and top 3, respectively, while all of these methods failed to rank the latter variant. Taken together, Berrylyzer demonstrated high efficiency for both autosomal dominant inheritance and autosomal recessive inheritance, significantly superior to existing tools.

### Performance evaluation across diverse disease subgroups

Subsequently, we explored whether the performance of Berrylyzer could be affected by the complexity and variability of disease spectrum. Cases with diagnostic and potential variants were included for analysis, and they were divided into six subgroups based on the clinical descriptions and the official HPO hierarchy, including abnormality of musculoskeletal system, abnormality of neurologic system, abnormality of cardiovascular system, abnormality of genitourinary system, abnormality of multiple systems, and abnormality of other systems (Figure 4 A). Cases presenting abnormalities of at least two systems were classified into subgroup of abnormality of multiple systems, and cases with limited number were combined into the subgroup of abnormality of other systems. As seen in Figure 4B and C, Berrylyzer consistently demonstrated high performance across different disease subgroups, with up to 80% of variants ranking within top 20 in almost all disease groups. For example, in the subgroup of abnormality of cardiovascular system, Berrylyzer reached a top 1 accuracy of 55.00% in the TSFMIH dataset, outperforming the second-best method at 10.00%. Similarly, in the subgroup of abnormality of neurologic system, Berrylyzer achieved a top 1 accuracy of 40.48% in the TTAHZU dataset, representing an increase of 8.93% over the second-best method Exomiser. However, performance disparities were observed in the subgroups of abnormality of genitourinary system, where Berrylyzer underperformed relative to Exomiser and PhenIX, highlighting context-specific limitations that warrant careful consideration in clinical application. Fetal structural abnormalities are highly etiological heterogeneity, for example cardiac malformation may result from hundreds of genetic causes. Berrylyzer’s high efficiency in prioritizing variants across the disease spectrum is critical beneficial for accurate prenatal diagnosis, especially in regions with limited medical resource.

**Figure 4.**
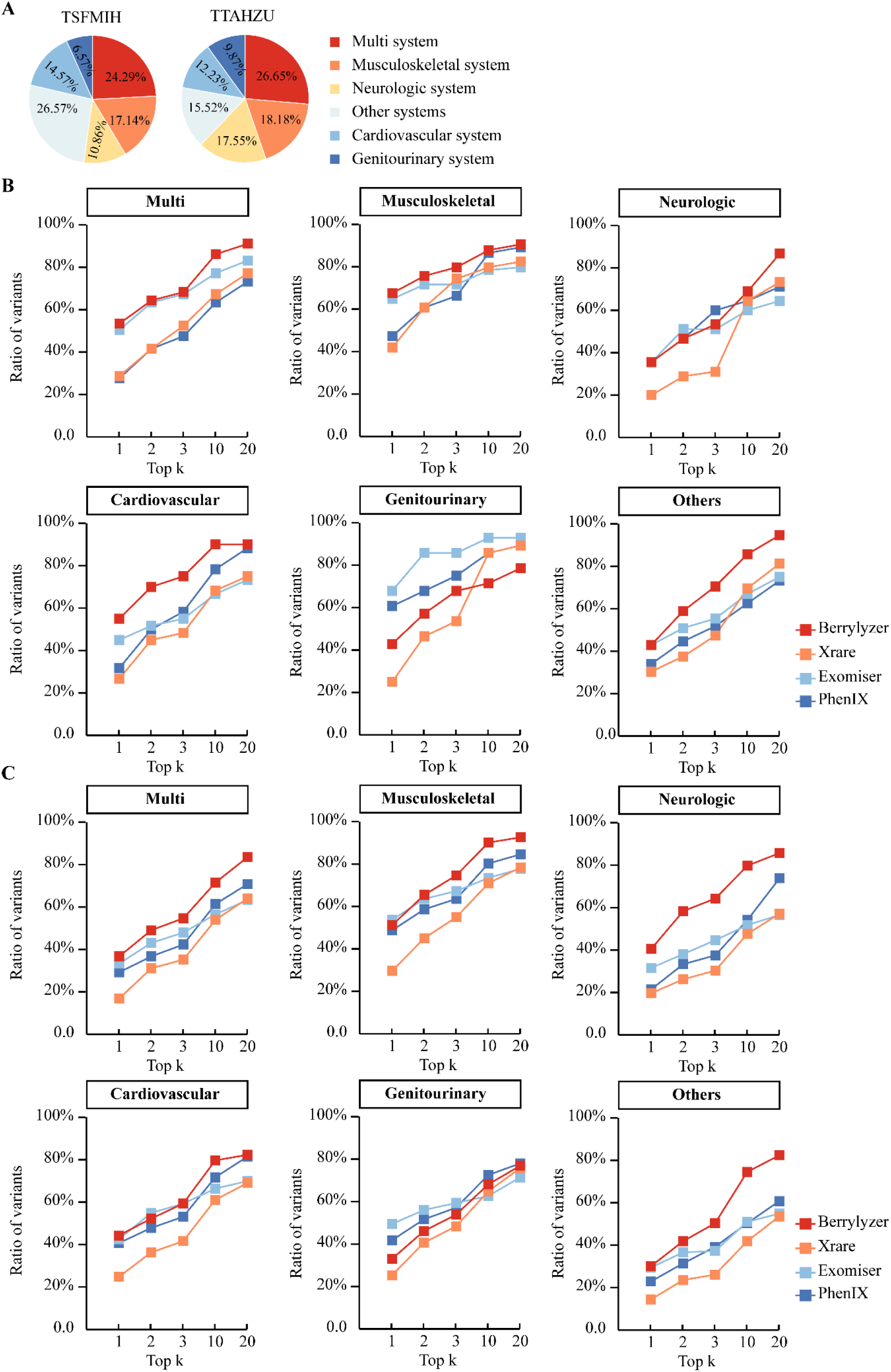
Performance evaluation across diverse disease subgroups. (A) The proportion of participants displaying diverse disease subgroups. (B-C) Distribution plots of performance on the TSFMIH cohort (B) and the TTAHZU cohort (C).

### Performance evaluation under different analytical strategies

In clinical practice, prenatal diagnosis often is performed with trio clinical sequencing, which enhances diagnostic accuracy. Here, we investigated the performance of Berrylyzer under different analysis strategies, namely “proband only” strategy and “trio” strategy. The results showed that, Berrylyzer exhibited comparably to, or even better performance than the three benchmarking methods, confirming its robustness in diverse clinical situations. Specifically, in “proband only” strategy, it achieved a top 20 accuracy of 90.00% on the TSFMIH cohort (Figure 5 A) and and 88.89% on the TTAHZU cohort (Figure 5 B), respectively. Similarly, Berrylyzer maintained its performance superiority in the “trio” strategy, with correctly ranking 89.55% of variants within top 20 on the TSFMIH cohort and 82.76% on the TTAHZU cohort. These results confirm Berrylyzer’s flexibility in adapting to different clinical workflows, which is essential for its application in routine prenatal diagnostic practice.

**Figure 5.**
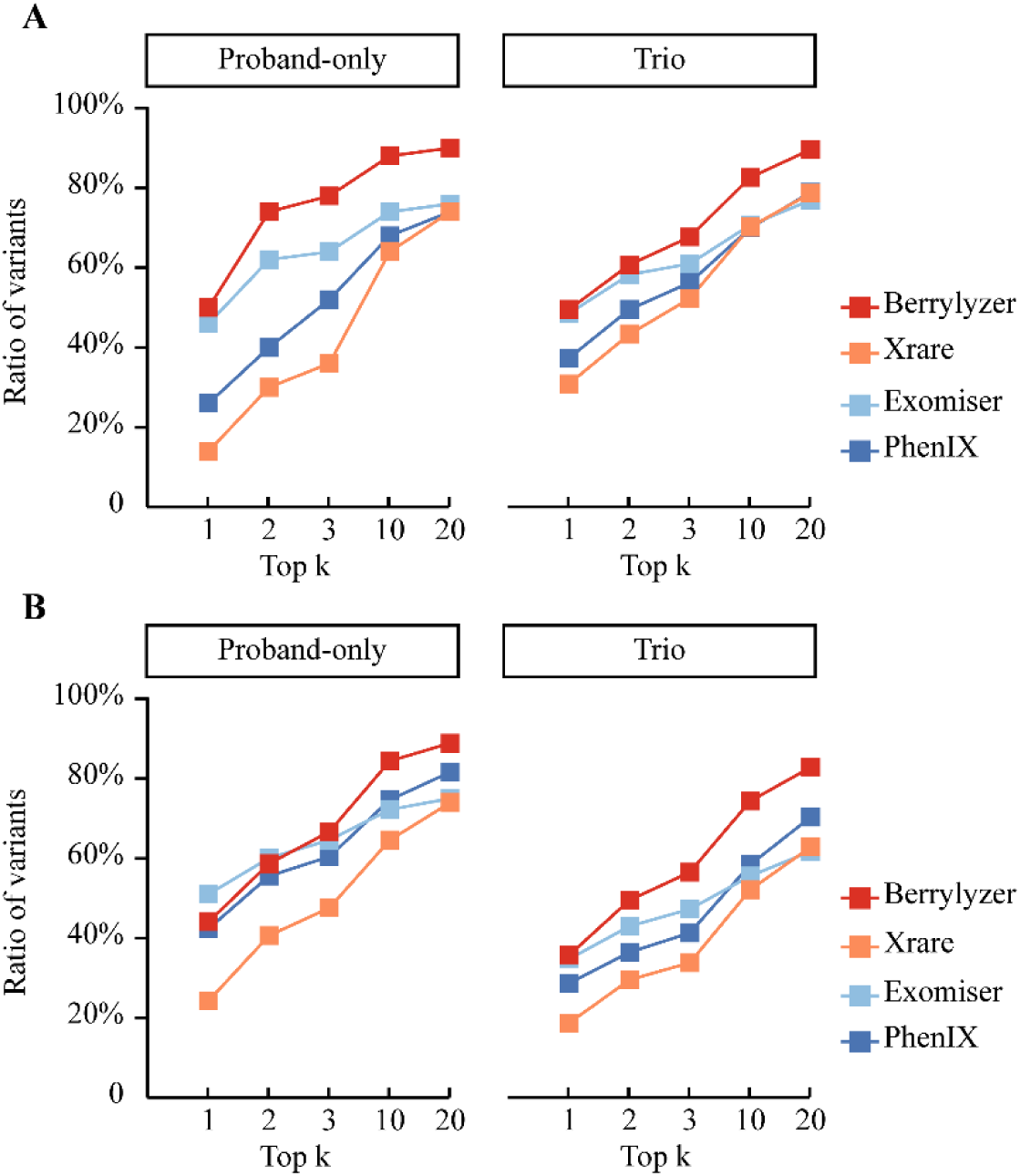
Performance evaluation across different analytical strategies. Distribution plots of performance on the TSFMIH cohort (A) and the TTAHZU cohort (B).

### Performance analysis with free-text clinical description and HPO

To comprehensively evaluate the performance of Berrylyzer, we assessed its accuracy for variant prioritization under two distinct input modalities: (1) free-text clinical descriptions and (2) standardized HPO terms. This evaluation was conducted on a curated subset of 709 cases from the TTAHZU cohort, who had been assigned HPO annotations during routine interpretation. As shown in Supplementary Figure 1, Berrylyzer demonstrated consistently high and comparable performance across both input formats: when provide with free-text clinical descriptions, the proportions of variants ranked within top 1, top 2, top 3, top 10, and top 20 were 39.93%, 54.76%, 62.00%, 76.56% and 85.71%, respectively; with HPO terms, the corresponding values were 37.36%, 51.56%, 58.97%, 76.56%, and 84.16%. These results highlight Berrylyzer’s robustness and reliability as a fully automated variant interpretation system, supporting efficient and accessible prenatal genetic diagnosis.

### Performance extension for secondary finding

Occasionally, prenatal genomic sequencing identifies secondary findings which are unrelated to the primary indication for testing but also have potential clinical significance (27). Thus, in addition to the concentration on the primary findings which explained the patients’ phenotypes, we also explored whether Berrylyzer could effectively prioritize the secondary findings. Our results revealed notable difference in performance among the four methods. On the TSFMIH cohort, Berrylyzer achieved a top 20 accuracy of 87.91%, which was slightly higher than that of Xrare (84.62%) and significantly exceeded the performance of Exomiser (61.54%) and PhenIX (67.58%) (Supplementary Figure 2A). However, Xrare exhibited the best performance on the TTAHZU cohort with a top 20 accuracy of 88.33% (Supplementary Figure 2B), surpassing the second best method Berrylyzer (81.67%). This result was contrast to the comparatively poor performance of Xrare observed in other clinical scenarios in this study. These results demonstrate that Berrylyzer maintains competitive efficacy in detecting secondary findings, further underscoring its versatility and potential utility in routine prenatal genomic analysis.

## Discussion

In this study, we present Berrylyzer, a novel, end-to-end computational framework to address the key clinical and technological challenges faced in variant interpretation during prenatal diagnosis of human Mendelian disorders. Through rigorous benchmarking across two independent, real-world prenatal cohorts, we demonstrated that Berrylyzer consistently outperformed existing prioritization tools, including Xrare, Exomiser, and PhenIX on both ranking yield and accuracy. Stratified analysis across disease categories, inheritance patterns, analytical strategies, and phenotypic input modalities further confirmed its robustness and broad clinical utility, even in complex clinical scenarios. To the best of our knowledge, this work represents the pioneer to establish an AI-driven variant prioritization framework in the field of prenatal diagnosis, making a paradigm shift from labor-intensive, manual curation toward intelligent, precision-guided clinical decision support.

Berrylyzer introduces several core innovations to address the inherent bottlenecks in prenatal variant interpretation: (1) time-sensitive computational design. Prenatal clinical decision-making is constrained by narrow temporal windows spanning only days to weeks, which requires rapid diagnostic turnaround. Conventional analytical workflows, which heavily rely on manual interpretation of large-scale genomic data, impose significant delays. Benchmarking results indicate that Berrylyzer correctly ranked over 50% of diagnostic variants first, enabling definitive diagnosis after reviewing a single candidate variant, thereby greatly induces the turnaround time. Notably, superior to existing state-of-the-art tools, Berrylyzer eliminates format constraints on phenotypic input by integrating NLP technology capable of parsing unstructured phenotypic information, achieving a fully automated, end-to-end prioritization framework. This ensures seamless integration with laboratory information systems and electronic health records, supporting scalable deployment across diverse clinical environments; (2) comprehensive prenatal phenotype dictionary. Current disease databases remain marked underrepresentation of prenatal manifestations, limiting the sensitivity of phenotype-driven prioritization in this context. To bridge this gap, we curated a prenatal phenotype dictionary. This resource enables comprehensive recognition of fetal phenotypes and further supports precise, ontology-aligned mapping between observed prenatal phenotypes and candidate Mendelian disorders, significantly improving the fidelity of phenotype-driven prioritization in prenatal scenarios compared to existing tools; (3) clinician-centered interpretability and decision transparency. Given the high-stakes nature of prenatal decision-making, analytical interpretability and clinical controllability are particularly essential for variant prioritization tools. Berrylyzer employs a hierarchical evidence presentation architecture: on one hand, it supports interactive keywords refinement to enhance the accuracy of phenotypic concept extraction; on the other hand, it provides step-by-step scoring outputs annotated with relevant phenotypic and genomic evidence, thereby fostering clinician confidence. This “human-in-the-loop” preserves clinical oversight throughout each stage of the process while maintaining analytical rigor; and (4) lightweight deployment in clinical practice. Berrylyzer supports on-premises deployment to ensure full compliance with data privacy and security regulations. Recognizing the heterogeneity of clinical computing environments, particularly in resource-constrained or edge settings, Berrylyzer employs lightweight architecture optimized for low memory consumption. While higher than Exomiser (∼8 GB RAM for exome-scale analysis), Berrylyzer’s modest increase in computational demand is deliberately traded for substantially richer annotations which are clinically justified by gains in prioritization sensitivity and yield. Future iterations will pursue further efficiency gains without compromising the annotation coverage essential to Berrylyzer’s clinical performance advantage. Moreover, the modular architecture further enhances clinical flexibility. The phenotypic scoring module enables preliminary fetal assessment, even in the absence of genomic data, thereby supporting early differential diagnosis or targeted disease screening. Besides, given the dynamic evolution of fetal phenotypes across gestation, reanalysis is often warranted upon the emergence of new or more specific clinical findings (28). Berrylyzer supports interactive reanalysis: clinicians can incrementally update, refine, or expand phenotypic inputs, which facilitates timely reanalysis especially in previously undiagnosed cases.

Despite Berrylyzer’s outstanding performance and broad clinical applicability, several limitations warrant further refinement. The current algorithm is specifically designed to rank SNVs and InDels, but do not support other clinically significant variant types, such as copy number variants and structural variants. Future work will extend the algorithm to be compatible with full spectrum of variant types, thereby enabling unified analysis of genomic data. Besides, Berrylyzer is designed explicitly for clinical diagnosis but not for research, which exclusively relies on well-established gene-disease associations in the disease databases. While this design ensures reliability of ranking results, it constrains the potential for identifying novel gene-disease correlations. Previous studies have demonstrated the diagnostic rate of WES for prenatal diagnosis of unselected fetal anomalies ranges from approximately 8.5% to 10% (29, 30). Given that the ongoing discoveries of novel gene-disease associations are expected to increase the diagnostic yield (31), Berrylyzer has established a periodic update mechanism ensuring synchronization between the underlying knowledge base and domain advancements, thus further supporting periodic reanalysis of unresolved cases. Moreover, rigorous validation across more independent clinical centers, diverse experimental platforms, and heterogeneous analytical workflows are required to confirm Berrylyzer’s generalizability. Concurrently, iterative self-training and algorithmic refinement grounded in longitudinal, real-world clinical data will contribute to scalable integration into practical diagnosis workflows.

## Conclusion

Systematic evaluation based on multi-center, real-world prenatal cohorts demonstrates that Berrylyzer delivers a robust, clinically deployable framework for variant prioritization in prenatal genetic diagnosis. Its high interpretability and lightweight architecture will jointly facilitate seamless integration into routine clinical workflows, thereby providing foundational technical infrastructure for the advancement of precision medicine in prenatal care. This study not only introduces a validated, AI-driven variant prioritization tool for clinical prenatal diagnosis but also establishes a methodological paradigm for the development of intelligent diagnostic system targeting Mendelian disorders.

## Abbreviations

ACMG: the American College of Medical Genetics and Genomics
AMP: the Association for Molecular Pathology
AI: Artificial intelligence
BERT: Bidirectional Encoder Representation from Transformers
CHPO: Chinese Human Phenotype Ontology
HPO: Human Phenotype Ontology
InDel: Insertion and Deletion
LLM: Large language model
MAF: Minor allele frequency
NER: Named entity recognition
NLP: Natural language processing
OMIM: the Online Mendelian Inheritance in Man
SNV: Single nucleotide variants
VCF: Variant call format
VUS: Variant of unknown significance
WE: Whole-exome sequencing

## Declarations

### Ethics approval and consent to participate

This study was approved by the Ethics Committee of the Shanghai First Maternity and Infant Hospital (KS25423). All participants in this study provided written informed consent.

### Consent for publication

Not applicable.

### Data availability

The data in the study are available from the corresponding author on reasonable request.

### Author contributions

G.Z. conceived the study and coordinated its overall implementation. X. Z., Y.T., K.S., N. L., P. Z., and N.F. recruited participants and performed clinical evaluations. N.Z. and S.L. collected and collated the datasets. M.M., L.L., Q.D., and X.L. analyzed the data and drafted the the initial manuscript. A.M. and Y.L. edited and revised the manuscript. All authors reviewed and approved the final version of the manuscript.

## Acknowledgements

Not applicable.

