## Supplementary Figure 1 and Supplementary Figure 2 for "Berrylyzer-an Efficient, Traceable, and Lightweight Intelligent Agentic System for Prenatal Genetic Diagnosis"

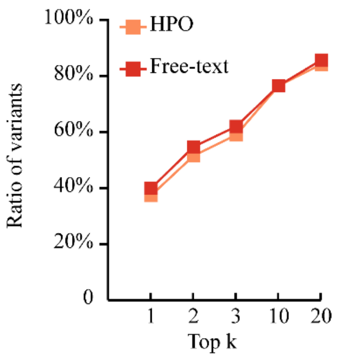


**Supplementary Figure 1**. Performance of Berrylyzer across diverse phenotypic inputs: free-text clinical descriptions and HPO terms.

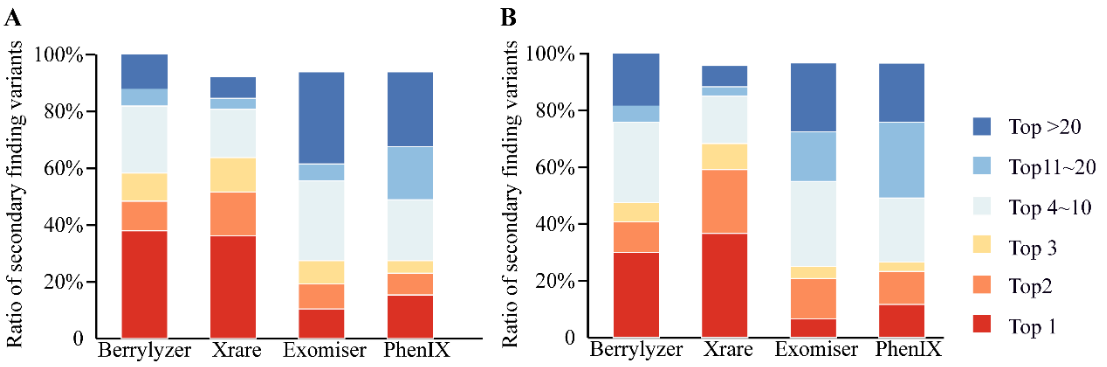


**Supplementary Figure 2.** Performance evaluation for secondary findings in the two cohorts. (A) TSFMIH cohort; (B) TTAHZU cohort.
